# Low serum 25-hydroxyvitamin D (25[OH]D) levels in patients hospitalised with COVID-19 are associated with greater disease severity: results of a local audit of practice

**DOI:** 10.1101/2020.06.21.20136903

**Authors:** Grigorios Panagiotou, Su Ann Tee, Yasir Ihsan, Waseem Athar, Gabriella Marchitelli, Donna Kelly, Christopher S. Boot, Nadia Stock, James Macfarlane, Adrian R. Martineau, Graham Burns, Richard Quinton

**Author notes:** contributed equally to this work (Grigorios Panagiotou, Su Ann Tee). **Corresponding author, reprint requests addressed to:** Dr Richard Quinton, Royal Victoria Infirmary, Newcastle upon Tyne, United Kingdom, NE1 4LP.

## Abstract

**Objectives:** To audit implementation of a local protocol for the treatment of vitamin D deficiency (VDD) among patients hospitalized for Coronavirus Disease 2019 (COVID-19), including an assessment of the prevalence of VDD in these patients, and of potential associations with disease severity and fatality.

**Design:** This was not a study or clinical trial, but rather a retrospective interim audit (Newcastle-upon-Tyne Hospitals Registration No. 10075) of a local clinical care pathway for hospitalized patients with COVID-19-related illness. The Information (Caldicott) Guardian permitted these data to be shared beyond the confines of our institution.

**Setting:** A large tertiary academic NHS Foundation Trust in the North East of England, UK, providing care to COVID-19 patients.

**Participants:** One hundred thirty-four hospitalized patients with documented COVID-19 infection.

**Main outcome measures:** Adherence to local investigation and treatment protocol; prevalence of VDD, and relationship of baseline serum 25(OH)D with markers of COVID-19 severity and inpatient fatality versus recovery.

**Results:** 55.8% of eligible patients received Colecalciferol replacement, albeit not always loaded as rapidly as our protocol suggested, and no cases of new hypercalcaemia occurred following treatment. Patients admitted to ITU were younger than those managed on medical wards (61.1 years ± 11.8 *vs*. 76.4 years ± 14.9, p<0.001), with greater prevalence of hypertension, and higher baseline respiratory rate, National Early Warning Score-2 and C-reactive protein level. While mean serum 25(OH)D levels were comparable [*i*.*e*. ITU: 33.5 nmol/L ± 16.8 *vs*. Non-ITU: 48.1 nmol/L ± 38.2, mean difference for Ln-transformed-25(OH)D: 0.14, 95% Confidence Interval (CI) (−0.15, 0.41), p=0.3], only 19% of ITU patients had 25(OH)D levels greater than 50 nmol/L *vs*. 39.1% of non-ITU patients (p=0.02). However, we found no association with fatality, potentially due to small sample size, limitations of no-trial data and, potentially, the prompt diagnosis and treatment of VDD.

**Conclusions:** Subject to the inherent limitations of observational (non-trial) audit data, analysed retrospectively, we found that patients requiring ITU admission were more frequently vitamin D deficient than those managed on medical wards, despite being significantly younger. Larger prospective studies and/or clinical trials are needed to elucidate the role of vitamin D as a preventive and/or therapeutic strategy for mitigating the effects of COVID-19 infection in patients with VDD.

**What is already known on this topic:**

- Vitamin D deficiency (VDD) is associated with increased risk for acute respiratory tract infections
- A link between VDD and severity of COVID-19 pathophysiology has been proposed
- Two recent (non-peer-reviewed) studies have reported crude associations between VDD in defined geographic populations and COVID-19 severity and mortality

**What this study adds:**

- *These data do not arise from a clinical study*; rather from an audit of a local replacement protocol for VDD in COVID-19 inpatients in a large UK centre, which found a significantly higher prevalence of VDD among ITU patients compared to non-ITU patients, despite the ITU patients being significantly younger.
- Prompt treatment of VDD following a local protocol did not result in any adverse events, such as hypercalcaemia.
- Whilst by no means conclusive, these data suggest an important association between VDD and COVID-19 severity; hence our report of interim findings in advance of achieving completed outcomes (fatality *vs*. recovery) for all patients.
- There is an urgent need for larger studies exploring vitamin D as a potential preventative measure and/or treatment of Covid-19-related illness among individuals with VDD.

## Introduction

The global pandemic of Coronavirus Disease 2019 (COVID-19) caused by Severe Acute Respiratory Syndrome Coronavirus-2 (SARS-CoV-2) poses serious medical and socio-economic challenges. A variety of puzzling epidemiological associations with fatality rates among infected individuals signpost major genetic and/or environmental predispositions. Higher fatality is strikingly associated with male sex [1], advancing age, obesity and metabolic syndrome, areas of high population density, climatic factors (low ambient temperature and high geographic latitude) and (in the UK and North America), the full spectrum of darker-skinned ethnicities [2].

Although classically considered as a fat-soluble vitamin, vitamin D3 is a photosynthesized pre-pro-hormone, whose biosynthetic pathway begins with solar UVB irradiation of 7-dehydrocholesterol when bare skin is exposed to strong sunlight, and exhibits multifaceted effects that extend beyond calcium and bone metabolism. Vitamin D receptors are ubiquitous in human tissues and are highly-expressed in B- and T-lymphocytes, suggesting a role in modulating innate and adaptive immune response [3]. Levels of 25-hydroxyvitamin D (the biologically most relevant metabolite to measure) reach their nadir at the end of winter and are associated with several adverse health outcomes, such as a higher risk for acute respiratory tract infections (ARTIs) during winter [4], which is mitigated by vitamin D supplementation [5].

Due to their northern latitude and prevailing cloud cover, the British Isles experience particularly low ambient UVB compared with continental Europe. Moreover, unlike our Nordic neighbours, intake of vitamin D-rich oily fish and vitamin D-containing dietary supplements is low and statutory fortification of foods is minimal. Thus, 50% of 45 year-old UK white males had VDD (<50nmol/L) at the end-winter nadir, with proportionately even lower levels in Scotland and Northern England [6]. Since then, the UK population has become more ethnically diverse and increasingly obese, with body fat tending to sequester vitamin D and thus lower serum levels. Indeed, several thousand cases of childhood rickets continue to be reported in the UK each year [7].

The recent outbreak of COVID-19 in the UK began in late winter and was associated with higher mortality in elderly and obese populations, as well individuals with darker skin; in all of whom VDD is more prominent [8 9]. COVID-19 had caused proportionately fewer fatalities in countries closer to the equator [10], suggesting a potential involvement of vitamin D in COVID-19 pathophysiology, and supplementation has therefore been recommended, despite the absence of direct evidence [11]. However, more recent data from UK biobank using historic levels, unrelated to season, did not find any association, but neither did they do so with well-established risk factors for fatality, hypertension and diabetes [12], which raises significant questions.

Therefore, two randomized controlled trials involving high-dose vitamin D supplementation in COVID-19 patients are currently underway (ClinicalTrials.gov identifier numbers: NCT04344041 and NCT04334005) [13 14], but may not report within the time-frame of this pandemic. However, as North East England has some of the highest prevalence of VDD [15], Physicians in Newcastle-upon-Tyne Hospitals (NuTH) NHS Foundation Trust decided - on the basis of available evidence - that the safest course of action was to measure serum 25(OH)D levels on admission in patients being treated for COVID-19, so as to inform a treatment protocol (summarized in Table 1) adjusted according to the severity of baseline deficiency.

**Table 1.** NuTH NHS Foundation Trust treatment protocol for vitamin D deficiency in COVID-19

| **25(OH)D level (nmol/L)** | **Dose of Colecalciferol prescribed** |
| --- | --- |
| Less than 13 | 300,000 international Units oral one-off dose<br>Followed by 1600 international Units oral daily |
| 13-25 | 200,000 international Units oral one-off dose<br>Followed by 800 international Units oral daily |
| 26-40 | 100,000 international Units oral one-off dose<br>Followed by 800 international Units oral daily |
| 41-74 | 800 international Units oral daily |
| Equal or greater than 75 | No replacement |

It was also decided to audit this local protocol as soon as practicable, in order to determine whether the data supported its continuation and whether there might be important lessons therein for a wider medical audience.

## METHODOLOGY OF LOCAL PROTOCOL AND AUDIT

Our local protocol (see Table 1) was implemented in mid-April 2020, along with the intention to audit the data as it accumulated (NuTH NHS Foundation Trust Clinical Governance & Audit Registration No. 10075, with permission from the Information Guardian to share results externally). Serum 25(OH)D levels were measured in 134 affected patients admitted to NuTH NHS Foundation Trust. In our institution, SARS-CoV-2-infected patients were defined by a positive swab [using reverse-transcriptase polymerase chain reaction (RT-PCR)], or – if swab-negative – by the presence of characteristic clinical, radiological and haematological findings of COVID-19 infection. Measurement of serum 25(OH)D was performed using the Roche Elecsys vitamin D total II immunoassay (Roche Diagnostics Ltd, Rotkreuz, Switzerland). In cases of undetectable levels of serum 25(OH)D, the lower limit of sensitivity of our assay (*i*.*e*. 8 nmol/L) divided by square root of 2 was used for analyses. A cut-off of greater than 50 nmol/L was defined as normal range as per previously published guidelines [16].

Patients found to have VDD were treated wherever possible according to the protocol given in Table 1, wherein the replacement dosage was determined by severity of deficiency (Table 1), based on pharmacokinetic data from Romagnoli, *et al*. [17]. Patients who did not receive treatment included some who had already been discharged home, were deceased, or were receiving “end-of-life care” by the time 25(OH)D results were available (n=50/113; 44.2%). No adverse effects, such as hypercalcemia, were reported after treatment.

For all patients, their clinical observations at presentation [including National Early Warning Score-2 (NEWS-2) score, initial heart rate, respiratory rate, blood pressure and temperature], initial markers of inflammatory response [C-reactive protein (CRP), procalcitonin] and haematology results [total White Cell Count (WCC); lymphocytes; eosinophils] were retrieved from electronic records.

Patients were managed according to disease severity, with the sickest patients admitted to Intensive Therapy Unit (ITU) and milder cases, or those with ward-based ceilings of care (*i*.*e*. trial of Continuous Positive Airway Pressure (CPAP) or Non-Invasive Ventilation (NIV) as maximal therapy) being managed on medical wards (“non-ITU group”). Final outcome of admission (where available) was recorded as discharge or death.

Data collection and methods employed herein were in accordance guidance from our Information Guardian and with the Declaration of Helsinki; aimed to the benefit of our patients. Data were anonymized during analysis and confidentiality of clinical data adhered to Caldicott Principles (NuTH Information Governance and Security Caldicott approval No. 7584). Nature of this analysis was observational as part of an audit project to aid clinicians in our institution abide to our internal protocol for vitamin D replacement during COVID-19 pandemic and, therefore, our data and manuscript were submitted to the Trust Clinical Ethics Advisory Group and approved for use in external publication.

## Statistical Analysis

Statistical analysis was performed with SPSS version 26.0 (IBM Corp., Armonk, NY), as appropriate. Data are presented as mean ± standard deviation (SD), unless stated otherwise. Normality of distribution was assessed with Kolmogorov-Smirnov test and variables including 25(OH)D and CRP levels as well as respiratory rate, WCC and lymphocyte count were logarithmically transformed for comparisons, if not normally distributed. Between group comparisons were assessed with independent t-test or Mann-Whitney U test in case of two groups, and Analysis of Variance and/or Kruskal-Wallis test in case of three or more groups. Associations between continuous variables were computed using Pearson’s or Spearman’s correlation coefficient and chi-square in case of categorical variables. Logistic regression models adjusting for age, gender, presence of co-morbidities and CRP levels were used to identify predictors of outcomes. Level of statistical significance was set at 0.05.

## Results

Descriptive characteristics of our audit population are summarized in Table 2. It was striking that, overall, 90/134 (66.4%) of Covid-19 inpatients had vitamin D insufficiency (<50 nmol/L); 50/134 (37.3%) inpatients had deficiency (<25 nmol/L) and 29/134 inpatients (21.6%) had very severe deficiency (≤15 nmol/L).

**Table 2.** Descriptive characteristics of audit participants.

|  | *Non- ITU wards*<br>(N=92) | *Intensive Therapy Unit*<br>(N=42) | *p- value* |
| --- | --- | --- | --- |
| Females (% of group subtotal) | 44 (47.8%) | 17 (39.5%) | 0.30 |
| Age (years) | 76.4 ± 14.9 | 61.1 ± 11.8 | **<0.001** |
| Ethnicity (N, %) |  |  |  |
| - Caucasian | 88 (95.7%) | 40 (95.2%) | 0.83 |
| - Asian | 3 (3.3%) | 1 (2.4%) |  |
| - Afro-Caribbean | 1 (1.1%) | 0 |  |
| - Other | 0 | 1 (2.4%) |  |
| Comorbidities (N, %) | N=79 | N=35 |  |
| - Hypertension | 32 (40.5%) | 24 (68.6%) | **<0.01** |
| - Diabetes | 24 (30.4%) | 14 (40%) | 0.27 |
| - Obesity | 5 (6.3%) | 9 (25.7%) | **<0.01** |
| - Malignancy | 12 (15.2%) | 3 (8.6%) | 0.36 |
| - Respiratory | 30 (38%) | 12 (34.3%) | 0.57 |
| - Cardiovascular disease | 15 (19%) | 5 (14.3%) | 0.59 |
| - Kidney and Liver diseases | 15 (19) | 4 (11.4%) | 0.35 |
| - Other | 11 (13.9%) | 3 (8.6%) | 0.48 |
| Systolic Blood pressure (mmHg) | 125.3 ± 21.1 | 120.2 ± 18.5 | 0.18 |
| Diastolic Blood pressure (mmHg) | 71.8 ± 12.4 | 68.8 ± 11.5 | 0.22 |
| Heart Rate (per min) | 90.2 ± 20.9 | 92.4 ± 20.0 | 0.54 |
| Respiratory Rate (per min) | 21.5 ± 5.1 | 24.8 ± 7.0 | **<0.01\*** |
| Body Temperature (°C) | 37.0 ± 0.9 | 37.5 ± 1.1 | **0.02** |
| O<sub>2</sub> saturation (%) | 93.1 ± 6.6 | 93.3 ± 4.7 | 0.77 |
| White Blood Cell Count | 8.9 ± 3.9 | 8.4 ± 3.8 | 0.63* |
| Lymphocyte Count | 0.1 ± 0.6 | 1.3 ± 1.6 | 0.20* |
| Eosinophil | 0.05 ± 0.11 | 0.03 ± 0.07 | 0.43 |
| C-Reactive protein (mg/mL) | 107.9 ± 92.0 | 143.4 ± 99.4 | **0.045\*** |
| Procalcitonin (ng/mL) | 0.7 ± 1.8 | 1.4 ± 3.1 | 0.90 |
| 25-hydroxyvitamin D (nmol/L) | 48.1 ± 38.2 | 33.5 ± 16.8 | 0.30* |
| Vitamin D status (N, %) |  |  |  |
| - <50 nmol/L | 56 (60.9%) | 34 (81%) | **0.02** |
| - ≥50 nmol/L | 36 (39.1%) | 8 (19%) |  |

Significance is highlighted in bold. \*: Ln-transformed for comparisons.

Patients admitted to ITU were significantly younger (ITU: 61.1 years ± 11.8 *vs*. non-ITU: 76.4 years ± 14.9, p<0.001) and more frequently hypertensive, and had higher respiratory rate and CRP levels at presentation than non-ITU patients (Table 2). Admission NEWS-2 score was higher in ITU patients vs. non-ITU patients (p=0.01). In the overall cohort as well as within each patient group, 25(OH)D levels were not associated with increased oxygen requirements, NEWS-2 score, increased likelihood of radiological findings of COVID-19, and/or presence of co-morbidities such as hypertension, diabetes, obesity, respiratory disease, cardiovascular disease, liver and/or kidney disease or other (p>0.05 for all). Baseline 25(OH)D levels and prior use of vitamin D supplements were not associated with CRP levels (p>0.05 for both).

Although VDD is classically defined as a disease of old age, ITU patients had lower 25(OH)D levels compared to non-ITU patients [*i*.*e*. ITU: 33.5 nmol/L ± 16.8 *vs*. Non-ITU: 48.1 nmol/L ± 38.2, mean difference for logarithmically-transformed-25(OH)D: 0.14, 95% Confidence Interval (CI) (−0.15, 0.41)], although this did not reach statistical significance (p=0.3) possibly due to limited sample size. Nevertheless, ITU patients exhibited a significantly higher prevalence of VDD (level <50 nmol/L). Indeed, only 19% of ITU patients were vitamin D replete compared to 39.1% of non-ITU patients (p=0.02) (Table 2).

Overall, 63/113 (55.8%) eligible patients received treatment. Of those, 33/63 patients (52.4%) were treated as per our protocol and the rest received lower doses of oral Colecalciferol. Outcome data (discharge from hospital or death) was available for 110/134 patients (82.1%) at the time of reporting. Of those, 94 (85.5%) patients were discharged, 16 (14.5%) died; and 24 are still receiving inpatient care.

Using a binary logistic regression model, 25(OH)D levels were not associated with mortality [95% Confidence Interval, CI 0.97 (0.42, 2.23), p=0.94]. Further adjustments for potential covariates including age, gender, presence of comorbidities and CRP levels did not materially affect these results.

## Discussion

Increased mortality from COVID-19 is caused by severe acute respiratory syndrome, with cytokine storm and diffuse micro- and macrovascular thrombosis, with numerous treatments targeting these pathophysiological changes are currently being trialed. Due to the lack of a vaccine, or any robustly evidence-based drug treatment for this novel disease, it becomes important to determine which factors potentially influence disease severity, and ultimate outcome, from the start of the patient journey. Vitamin D is known to regulate immune responses and alter cytokine downstream pathways [18 19], and treatment of VDD therefore represents a promising modality for mitigating fatality arising from COVID-19 infections, not to mention being safe and cheap.

The UK’s Scientific Advisory Committee on Nutrition (SACN) previously recommended universal supplementation with 400 iU (10 ug) daily of vitamin D3, which was endorsed by PHE (Public Health England) in 2016. Nevertheless, current NHS-England guidance to primary care explicitly discourages prescribing maintenance therapy with vitamin D3 due to concerns about cost-effectiveness. Moreover, there has been no meaningful promotion to the general public (*e*.*g*. via lay media) of over-counter supplementation with the onset of winter - as is routine in Scandinavia - nor any commensurate diminution of sun-avoidance messages to the public by UK medical and lay media in spring and summer. Indeed, both UK government ministers and senior police officers repeatedly stated that sunbathing – as opposed to exercising – in public places during lock-down was unacceptable behaviour.

Vitamin D has been reported to lower the risk of respiratory tract infections through three mechanisms: maintaining tight junctions, killing enveloped viruses through induction of cathelicidin and defensins, and reducing production of pro-inflammatory cytokines by the innate immune system, thereby decreasing the risk of a cytokine storm [11]. Moreover, in relation to coronavirus, severity of bovine infections was associated with lower vitamin D levels [20]. Among the previously proposed treatments for COVID-19, hydroxychloroquine has been shown to increase 25(OH)D levels [21], while vitamin D replacement therapy has been suggested as a simpler and less expensive alternative to tocilizumab, an anti-Interleukin-6 (IL-6) receptor monoclonal antibody [22].

A number of sociological, lifestyle-behavioral and genetic hypotheses have been proposed to explain the striking fatality among ethnic darker-skinned residents of the UK and North America, including healthcare workers. A major problem with any genetically-based hypothesis of vulnerability is that differences between these individual affected ethnicities (African, South Asian, East Asian, Southeast Asian, Hispano-American, Middle Eastern, *etc*) are likely greater than between each individual group and the generality of Caucasians. Moreover, the Indian state of Kerala has recorded just 500 COVID-19 infections and 4 deaths [23], which is possibly less than the number of healthcare workers of Keralan origin who have died of COVID-19 in the UK. Finally, although socio-economic arguments are compelling, the almost exclusively ethnic minority roll-call of COVID-19-related deaths among doctors and dentists in the UK (18 of 19 deaths) [24] cannot realistically be ascribed to deprivation. Our inpatient cohort was almost entirely Caucasian, reflecting the demographic make-up of our region, and thus did not directly add to this issue.

Due to the ongoing pandemic, data on vitamin D status in COVID-19 patients so far has been patchy. To the best of our knowledge, no such studies have been published regarding UK or European inpatient cohorts with COVID-19. A recent publication highlighted a potential crude association between 25(OH)D levels and COVID-19 mortality, in that countries with increased mortality from COVID-19 also exhibit a high prevalence of VDD within the general population [25]. Another study from Indonesia recruiting 780 individuals, showed that low pre-admission 25(OH)D levels were significantly associated with fatality from COVID-19 [26]. We did not find any significant association between vitamin D status and inpatient mortality, but this was not unexpected given that we had proactively treated all COVID-19 patients with VDD wherever possible. However, longer-term follow-up of these patients (which is ongoing), may yet reveal future differences in 28- or 90-day mortality.

Other groups have looked specifically at possible associations between vitamin D status and disease severity, defined as presence of adverse clinical features in some studies *(e*.*g*. increased respiratory rate, hypoxia), or in others, by need for ITU treatment. A retrospective analysis of pre-hospital vitamin D levels of patients followed up in two tertiary centers in South Asia suggested that COVID-19 patients with severe disease had lower serum 25(OH)D concentrations, compared to individuals with mild symptoms; though level of statistical significance is not provided [27]. A separate study recruiting individuals from Southeast Asia suggested low serum 25(OH)D was correlated with adverse disease outcomes [28].

CRP levels have also been used as a surrogate marker of COVID-19 severity. One such study, using assumption models with previously reported data on the association between vitamin D and CRP levels, concluded that VDD may be related to COVID-19 severity [29]. Indeed, we found that CRP levels were significantly higher in ITU *vs*. non-ITU patients, thus supporting the finding that hypovitaminosis D is associated with more severe COVID-19 infection.

In a small US study recruiting 20 patients, vitamin D insufficiency was evident in 11 out of 13 (84.6%) ITU patients compared to 57.1% of patients treated in standard medical wards [30]. The authors failed to reach statistical significance, but overall, these results concord with our observations. More importantly, the authors reported that all ITU patients aged less than 75 years had substantially low vitamin D levels. In our inpatient COVID-19 population, we found that only 19% of our ITU patients were vitamin D replete, which is startling, as this group was significantly younger and had fewer of the co-morbidities traditionally associated with VDD. This finding challenges previous beliefs that VDD is a problem limited to the elderly frail population, and has significant implications for influencing public health advice, especially in light of recent limited sun exposure for most individuals, resulting from lockdown measures to limit COVID-19 spread.

Our findings also suggest that, outwith the established risk factors thus far associated with COVID-19 mortality (age, gender and relevant co-morbidities); VDD may be an important and hitherto under-recognized determinant of illness severity.

Strengths of the present data include its novelty and assessment of serum 25(OH)D levels during admission of patients with COVID-19, followed by prompt treatment of any deficiency where possible. Limitations of these data include the small sample size and the observational nature of this audit. We also acknowledge that the cross-sectional analysis employed herein does not allow causality to be established, particularly given data suggesting that 25OHD may be a negative acute phase reactant due to dysregulated vitamin D metabolism in respiratory disease [31]. Nevertheless, these data provide an important impetus to the commissioning, design and interpretation of ongoing or future multi-center randomized controlled trials, which could conclusively define a therapeutic role of vitamin D in COVID-19.

In addition, our population consisted of predominantly Caucasian patients with moderate to severe symptoms from COVID-19 and, given that the vast majority of COVID-19 cases are asymptomatic or mildly symptomatic (not requiring hospital admission), community-based studies are needed to address whether adequate vitamin D concentrations reduce the risk for hospital attendance. We have proactively treated VDD in our patients per our Trust protocol described herein and this might partly explain the lack of association of baseline serum 25(OH)D with fatality. Also, since data collection was performed in a retrospective manner, patients included in this audit may have been actively treated with various other medications as part of clinical research trials to identify successful treatment for COVID-19 and it is therefore almost impossible to adjust for this in the analysis.

In conclusion, to the best of our knowledge this is the first report exploring serum 25(OH)D levels in a cohort of inpatients with COVID-19 in Europe. We have assessed serum 25(OH)D levels during acute admission for COVID-19 and found that VDD is significantly more prevalent among patients requiring admission to ITU, and thus necessarily associated with more severe COVID-19 disease. Vitamin D status was not associated with inpatient fatality in our cohort, possibly due to the limited sample size and our protocol for rapid diagnosis and treatment of VDD. Larger studies are urgently needed to further establish the role of vitamin D status in COVID-19 and, more importantly, explore the use of vitamin D as a potential therapeutic strategy to mitigate against this major new infection.

## Data Availability

The data that support the findings of this study are available from the corresponding author (RQ), upon reasonable request.

## Author contribution

GP, ST, YI and RQ conceived the audit project. ST and YI designed the data collection proforma. ST, YI, WA, GM, and DK collected data. CSB provided biochemistry input and advice. NS, JM, GB and RQ provided clinical care and supervised the audit. GP and ST interpreted data and drafted the manuscript. ARM and RQ interpreted data and revised the manuscript for important intellectual content. All authors approved the final version of the manuscript. RQ is the guarantor of this work and attests that all listed authors meet authorship criteria and that no others meeting the criteria have been omitted.

